# Seroepidemiology of SARS-CoV-2 infections in an urban Nicaraguan population

**DOI:** 10.1101/2021.02.25.21252447

**Authors:** Fredman González, Nadja A. Vielot, Michael Sciaudone, Christian Toval-Ruíz, Lakshmanane Premkumar, Lester Gutierrez, Edwing Centeno Cuadra, Patricia Blandón, Aravinda M. de Silva, Rebecca Rubinstein, Natalie Bowman, Sylvia Becker-Dreps, Filemon Bucardo

## Abstract

In a Nicaraguan population-based cohort, SARS-CoV-2 seroprevalence was 34%, with higher prevalence in children compared to adults. Having a seropositive household member was associated with a two-fold probability of individual seropositivity, suggesting a role for household transmission. Co-morbidities and preventive behaviors were not associated with SARS-CoV-2 seroprevalence.

## INTRODUCTION

SARS-CoV-2 transmission continues globally.^1^ The pandemic is now entering a prolonged phase, potentially causing multiple waves of infection in populations that have not attained community immunity. In this new phase, information on SARS-CoV-2 seroprevalence in different populations is urgently needed to understand the magnitude of SARS-CoV-2 spread in previous waves, predict future waves, and measure the risk of re-infection in previously exposed persons. Furthermore, little is known about SARS-CoV-2 seroprevalence in different age groups or in individuals with factors which place them at higher risk for poor outcomes.

Prior studies have measured SARS-CoV-2 seroprevalence in hotspots that were heavily impacted by infections in the first wave of the pandemic (Dec 2019-May 2020). In Wuhan, China, where the pandemic originated, seroprevalence was 4% in May 2020; in Lombardy, Italy, seroprevalence among blood donors reached 23% by April 2020.^2,3^ This wide range of estimates suggests that geographical, social, and economic differences, as well as the implementation of containment measures can greatly affect exposure to and infection with SARS-CoV-2.

Even less is known in low-and middle-income countries (LMICs), which have limited resources to perform molecular detection of SARS-CoV-2 in real time, complicating estimates of infection incidence and individual disease risk. Furthermore, LMICs have fewer resources to support remote work and schooling, hygiene measures, and vaccines, and households are often multi-generational. The goal of this study was to estimate the seroprevalence of SARS-CoV-2 in a population-based sample in León, Nicaragua since SARS-CoV-2 infections were first reported in March 2020. These data on the prevalence of past infections can be used to guide public health recommendations and inform the need for continuation of SARS-CoV-2 prevention measures.

## METHODS

### Study Design and Population

The Sapovirus-Associated Gastro-Enteritis (SAGE) study is a population-based birth cohort study in León, Nicaragua, described previously.^4^ This cohort provided a platform to access a sample of all ages to understand the seroprevalence of SARS-CoV-2 in a Latin American context, and to examine differences in participant characteristics by evidence of prior infection. The study population included high-income families in the city center and low-income families in peri-urban neighborhoods, creating a scientifically-informative gradient to evaluate socioeconomic and environmental risk factors for SARS-CoV-2 infection. Starting in July 2020, we contacted the household members of cohort children (both adults and children) and offered participation in this study. We also offered participation in the study for cohort children, including those who had reached 36 months of age. The study was approved by the Institutional Review Boards of the National Autonomous University of Nicaragua, León (UNAN-León, acta No. 170, 2020) and the University of North Carolina at Chapel Hill (Study #: 20-2126). All adult participants provided informed consent, and parental permission was required for children’s participation in the study.

In September and October 2020, we collected baseline demographic and health history data and collected blood from all participants for baseline SARS-CoV-2 serology. The current report summarizes the seroprevalence of SARS-CoV-2, stratifying by co-morbidities and sociodemographic factors.

### Specimen Collection and Laboratory Methods

SARS-CoV-2 infection induces the production of antibodies (Ab) against the spike protein and nucleocapsid protein, with most patients seroconverting within 2 weeks of symptom onset.^5^ We used an in-house enzyme-linked immunosorbent (ELISA) assay to measure antibodies (IgG, IgA, and IgM) to the receptor binding domain (RBD) of the SARS-CoV-2 spike protein, which we have previously shown to be highly sensitive and specific for detecting antibodies for at least six months among individuals experiencing symptomatic infection.^5^ The spike RBD-based assay does not cross-react with common endemic coronaviruses, and the magnitude of RDB antibody titers correlate with neutralizing antibody titers, currently the best correlate of protection against infection.^5^ In brief, ELISA plates (Greiner Bio-One #655061) were coated with 4µg/ml of the RBD antigen. Heat-inactivated serum diluted at 1:40 was subsequently added and alkaline phosphate conjugated secondary goat anti-human Abs (anti-IgG [Sigma], anti-IgA [Ab cam], and anti-IgM [Sigma]) were added at 1:2,500 dilution for detection. The immunologic reaction was developed with para-Nitrophenyl phosphate substrate (SIGMA). The optical density (OD) was measured after 15 min at 405nm. A serum was considered positive if the positive/negative (P/N) ratio between the serum OD and the negative-control OD was ≥2.57, to ensure 99.5% specificity per CDC guidelines.^6^

### Statistical Analysis

We analyzed cross-sectional data using frequencies and percentages to characterize SARS-CoV-2 seroprevalence, stratified by sex, age group, smoking status, presence of comorbidities, and socioeconomic characteristics of the household. We implemented generalized estimating equations to estimate prevalence ratios (PRs) comparing seropositivity proportion by select individual and household characteristics, accounting for clustering of individuals within households.

## RESULTS

Between September and October 2020, we enrolled 1,847 participants from 295 households. Of 1,351 individuals who provided serum samples, 456 were seropositive for SARS-CoV-2 (34%). In 192 households (65%), at least one household member had SARS-CoV-2 antibodies at the time of enrollment. In these households, the median number of seropositive members was 2 (interquartile range: 1, 3), and the maximum number was 9.

Women were less likely to be seropositive than men (PR: 0.90, 95% CI: 0.77, 1.06), and younger age groups were more likely to be seropositive than older age groups (PR: 0.93, 95% CI: 0.88, 0.99), with approximately half of seropositive individuals younger than 15 years (Table). Smoking and the presence of comorbidities was associated with a lower prevalence of seropositivity, though several of these associations are imprecise due to small numbers. Seropositivity was not associated with physical distancing or masking behavior, nor with economic status of the household. Seropositivity was twice as high among individuals who lived with another seropositive household member (PR: 1.97, 95% CI: 1.43, 2.69).

**Table.** Characteristics associated with SARS-CoV-2 seropositivity of individuals in León, Nicaragua.

|  | Seronegative<br>(n=895) | Seropositive<br>(n=456) | Prevalence Ratio<br>(95% Confidence Interval) |
| --- | --- | --- | --- |
| Individual and household characteristics | n (%) | n (%) |  |
| Sex |  |  |  |
| Male | 350 (63.8) | 199 (36.2) | 0.90 (0.77, 1.06) |
| Female | 545 (68.0) | 257 (32.0) |  |
| Age group in years (n=1,325) |  |  |  |
| 0-4 | 218 (64.3) | 121 (35.7) | 0.93 (0.88, 0.99) |
| 5-14 | 145 (59.4) | 99 (40.6) |  |
| 15-29 | 263 (68.0) | 124 (32.0) |  |
| 30-49 | 165 (72.7) | 62 (27.3) |  |
| 50-69 | 81 (73.0) | 30 (27.0) |  |
| 70+ | 10 (52.6) | 9 (47.4) |  |
| Smoking status (n=782)<sup>a</sup> |  |  |  |
| Ever smoker | 75 (72.8) | 28 (27.2) | 0.85 (0.62, 1.18) |
| Never smoker | 465 (68.5) | 214 (31.5) |  |
| Presence of comorbidities (n=1,312)<sup>b</sup> |  |  |  |
| No | 788 (66.6) | 396 (33.4) | 0.88 (0.67, 1.15) |
| Yes | 88 (68.8) | 40 (31.2) |  |
| Diabetes (n=128) |  |  |  |
| No | 68 (70.1) | 29 (29.9) | 1.12 (0.62, 2.01) |
| Yes | 20 (64.5) | 11 (35.5) |  |
| Hypertension (n=128) |  |  |  |
| No | 41 (66.1) | 21 (33.9) | 0.85 (0.51, 1.44) |
| Yes | 47 (71.2) | 19 (28.8) |  |
| Malignancy (n=128) |  |  |  |
| No | 86 (68.8) | 39 (31.2) | 1.20 (0.28, 5.01) |
| Yes | 2 (66.7) | 1 (33.3) |  |
| Autoimmune disease (n=128) |  |  |  |
| No | 86 (68.3) | 40 (31.7) | n/a |
| Yes | 2 (100.0) | 0 (0.0) |  |
| Physical distancing and masking behavior (n=1,325)<sup>c</sup> |  |  |  |
| Strict | 797 (66.7) | 398 (33.3) | 1.06 (0.91, 1.22) |
| Moderate | 31 (56.4) | 24 (43.6) |  |
| None | 49 (65.3) | 26 (34.7) |  |
| Crowding index (n=1,212) |  |  |  |
| Fewer than 2.7 people per bedroom | 535 (66.1) | 274 (33.9) | 0.91 (0.73, 1.15) |
| 2.7 or more people per bedroom | 277 (68.7) | 126 (31.3) |  |
| Poverty index (n=1,212)<sup>d</sup> |  |  |  |
| Not poor | 378 (67.0) | 186 (33.0) | 0.96 (0.82, 1.11) |
| Poor | 349 (65.6) | 183 (34.4) |  |
| Extremely poor | 85 (73.3) | 31 (26.7) |  |
| Household member is seropositive |  |  |  |
| No | 323 (36.1) | 74 (16.2) |  |
| Yes | 572 (63.9) | 382 (83.8) | 1.97 (1.43, 2.69) |

<sup>a</sup>Smoking status assessed among participants older than 13 years of age.
<sup>b</sup>Diagnosed by medical professional as reported by participant, including diabetes, hypertension, obesity, malignancy, and autoimmune disease.
<sup>c</sup>Strict = never left the house since first cases reported or occasionally leaves house and always uses a mask;
Moderate=Occasionally leaves the house and sometimes uses a mask; None = Occasionally leaves the house and never uses a mask or no distancing or quarantine behavior.
<sup>d</sup>Poverty index calculated based on presence of basic needs, including household sanitation, education, economic dependency, and household crowding.

## DISCUSSION

Our results show a high rate of seropositivity for SARS-CoV-2 antibodies in a Nicaraguan population-based cohort. This suggests the actual number of SARS-CoV-2 infections in Nicaragua was higher than official reports, which might be explained by a high rate of asymptomatic and mild cases that did not seek medical attention, or by limited testing of patients who received medical attention. Our seroprevalence results are similar to those of another study from Brazil,^7^ and those reported from SARS-CoV-2 hot spots in high income countries, such as Massachusetts and Northern Italy, although the Massachusetts study was not carried out in a population-based cohort.^2,8^ In these studies, the seroprevalence of SARS-CoV-2 antibodies was higher than expected based on the reported incidence of symptomatic SARS-CoV-2 infections in the area. It is possible we slightly underestimated the actual number of SARS-CoV-2 cases because of the high P/N cutoff used to ensure high assay specificity. A recent study has shown anti-SARS-CoV-2 spike protein IgG remains detectable in most cases for at least 6 months after symptomatic infection, and our data were collected within 6 months of the first reported SARS-CoV-2 case in Nicaragua.^9^ A recent cross-sectional study of Nicaraguan health care workers (HCWs) conducted over a one-month period found that 30% of participants had active SARS-CoV-2 infection, indicating that seroprevalence among Nicaraguan health care workers is likely higher than in the general population.^10^ About half of SARS-CoV-2 infected HCWs were asymptomatic, and, similar to our results, men were more likely to have been infected.^9^ Use of personal protective equipment is critical among HCWs, both to prevent contracting SARS-CoV-2 from infected patients and to prevent transmitting SARS-CoV-2 to patients, particularly when the HCW is asymptomatic.

We identified high seroprevalence among younger age groups, particularly in children between 5 and 14 years of age (42%). Age-based differences in seropositivity were observed in Brazil and Italy,^2,7^ however, they were more pronounced in our population. High seropositivity in children might be due to continued operation of public schools throughout the epidemic. As children are more likely to experience mild symptoms than adults,^11^ this finding suggests that school-aged children may contribute to active transmission, and controlling infections in this sub-population is essential to reduce the spread of SARS-CoV-2. We also found that most individuals reported practicing strict physical distancing or masking behavior outside of the home, though this did not have a significant impact on the seroprevalence. Transmission within the home may have been more important.^12^ The median household size in our sample was 7 members, with a maximum of 26 members. Indeed, SARS-CoV-2 seropositivity was two-fold more frequent for individuals with a seropositive household member as compared to individuals who did not have a seropositive household member.

In conclusion, we found a high SARS-CoV-2 seroprevalence in this Nicaraguan population and confirmed that reported SARS-CoV-2 case counts underestimated the true number of infections. We also show that this population has not yet attained the theoretical community immunity threshold, and so continued containment measures are necessary. In the future, this population could be followed to understand the risk of recurrent infection, and repeated seroprevalence measurements could provide further information on transmission dynamics.

## Data Availability

Data are not publicly available. Data requests may be considered at the discretion of the senior authors.

## ACKNOWLEDGEMENTS

We greatly appreciate Yorling Picado, Nancy Corea Munguia, Franco Soto, and the SAGE field team for their important contributions to this study.

